# Towards a Transactional Understanding of Stress and Coping Among Nursing Students: A qualitative interview study

**DOI:** 10.64898/2026.10.01.26363847

**Authors:** Han Yan, Anthony John O’Brien, Victoria Shaw, Kat Vakavosaki

## Abstract

**Introduction:** Nursing students experience stress arising from demanding academic requirements and emotionally challenging clinical placements, which may affect wellbeing, learning, and programme retention. This study explored nursing students’ and educators’ perceptions of stress, its impacts, and coping experiences within undergraduate nursing education in New Zealand.

**Methods:** A qualitative descriptive study was conducted with six undergraduate nursing students and six nurse educators at a New Zealand university. Data were collected through semi-structured interviews and analysed using inductive reflexive thematic analysis, with interpretation informed by the Transactional Model of Stress and Coping.

**Results:** Three themes were identified: stress from multiple sources, stress shaping wellbeing and learning, and navigating stress through appraisal and coping. Stress arose from academic, clinical, personal, and social demands and affected students’ academic engagement, health, and relationships. Participants described varied responses to stress, including different appraisals of challenging situations and coping strategies such as peer support, self-care, and use of institutional resources.

**Conclusion:** Stress in nursing education reflects a dynamic interaction between individual appraisal, coping resources, and educational and interpersonal contexts. Although findings are limited to one institution, they highlight the importance of accessible, flexible, and culturally responsive support and of addressing modifiable educational stressors alongside strengthening students’ coping resources.

## Introduction

Stress is a significant and widespread concern with important implications for health and wellbeing across populations (World Health Organization, 2023). Among university students, stress is especially pronounced in nursing, a discipline characterised by intensive academic demands, emotionally and physically challenging clinical placements, and ongoing professional expectations (Welch, 2023). In New Zealand, over 86% of nursing students report moderate to excessive stress (New Zealand Nurses Organisation, 2023).

Stress among nursing students extends beyond temporary discomfort and may affect academic performance, clinical competence and long-term professional development (Labrague et al., 2017). When unmanaged, stress can contribute to anxiety, burnout and attrition, with implications for both student wellbeing and the future nursing workforce (Ching et al., 2020; Zheng et al., 2022). Previous research has identified multiple and often overlapping sources of stress among nursing students, including academic workload, examinations, fear of failure, clinical placements, patient-care responsibilities, relationships with clinical staff, financial pressures and competing personal responsibilities (Labrague, 2024; Labrague et al., 2017). Clinical learning environments may create additional stress because students are required to apply developing knowledge and skills while managing concerns about competence, patient safety and professional expectations (Ching et al., 2020; Welch, 2023).

Nursing students use a range of strategies to manage these demands, including peer and family support, problem-solving, physical activity, self-care and avoidance-based coping (Labrague et al., 2018; Rafati et al., 2017). However, students do not respond to similar stressors in the same way. Individual appraisal, previous experiences, perceived coping capacity and available support may influence whether demands are appraised as threating or challenging and whether they are experiences as overwhelming and manageable (Ching et al., 2020; Lazarus & Folkman, 1984). This suggests that understanding nursing student stress requires consideration not only of the stressors encountered, but also of how these demands are interpreted and responded to.

Although nursing student stress and coping have been widely investigated internationally, important gaps remain in understanding how students interpret stress within their educational contexts and how these experiences are perceived by the educators who support them. Existing literature has focused predominantly on negative outcomes, particularly impacts on performance and wellbeing, while there is growing recognition that stress may be experienced as both detrimental and adaptive (Aljohani et al., 2021; Lavoie-Tremblay et al., 2022). Positive or adaptive experiences of stress remain comparatively underexamined among nursing students. Educator perspectives are also less frequently examined alongside student accounts, despite educators’ central role in shaping academic and clinical learning environments. These gaps are particularly relevant in New Zealand, where research examining nursing students’ experiences of stress remains limited (Solomon et al., 2025).

In New Zealand, undergraduate nursing education is typically delivered through three-year pre-registration Bachelor of Nursing programmes that integrate university-based academic learning with supervised clinical practice (Nursing Council of New Zealand, 2026). Students therefore move repeatedly between academic and clinical learning environments throughout their programmes, balancing academic requirements with the expectations and responsibilities of clinical placement. This structure provides an important context for understanding nursing student stress, particularly when academic demands continue alongside periods of clinical practice.

Examining both student and educator perspectives may reveal differences in how stress is understood and supported. These processes have important implications for student mental health, wellbeing, learning and coping, as well as their preparation for managing the emotional demands of future nursing practice. This study therefore explores how nursing students experience and appraise stress, and how educators understand student stress, to provide a more comprehensive understanding of stress in New Zealand nursing education.

## 1. Aim

The aim of this study was to describe nursing students’ and educators’ perceptions of stress, impacts of stress, and coping strategies among undergraduate nursing students in New Zealand.

## 2. Methods

### 2.1 Qualitative approach and research paradigm

This study adopted an interpretivist paradigm, recognising that meanings are shaped through individuals’ experiences and social contexts. A qualitative approach was adopted to explore nursing students’ and educators’ experiences of stress within nursing education (Sandelowski, 2000). Data were analysed using inductive thematic analysis, allowing patterns and themes to be generated from participants’ accounts rather than being guided by predetermined coding frameworks (Clarke & Braun, 2017).

### 2.2 Theoretical orientation

The Transactional Model of Stress and Coping (Lazarus & Folkman, 1984) was used as a conceptual framework, conceptualising stress as a dynamic process shaped by primary appraisal of threat or challenge and secondary appraisal of coping capacity. In this study, the model offered a useful perspective for understanding nursing students’ and educators’ experiences of stress and coping across academic and clinical contexts (Figure 1).

**Figure 1:**
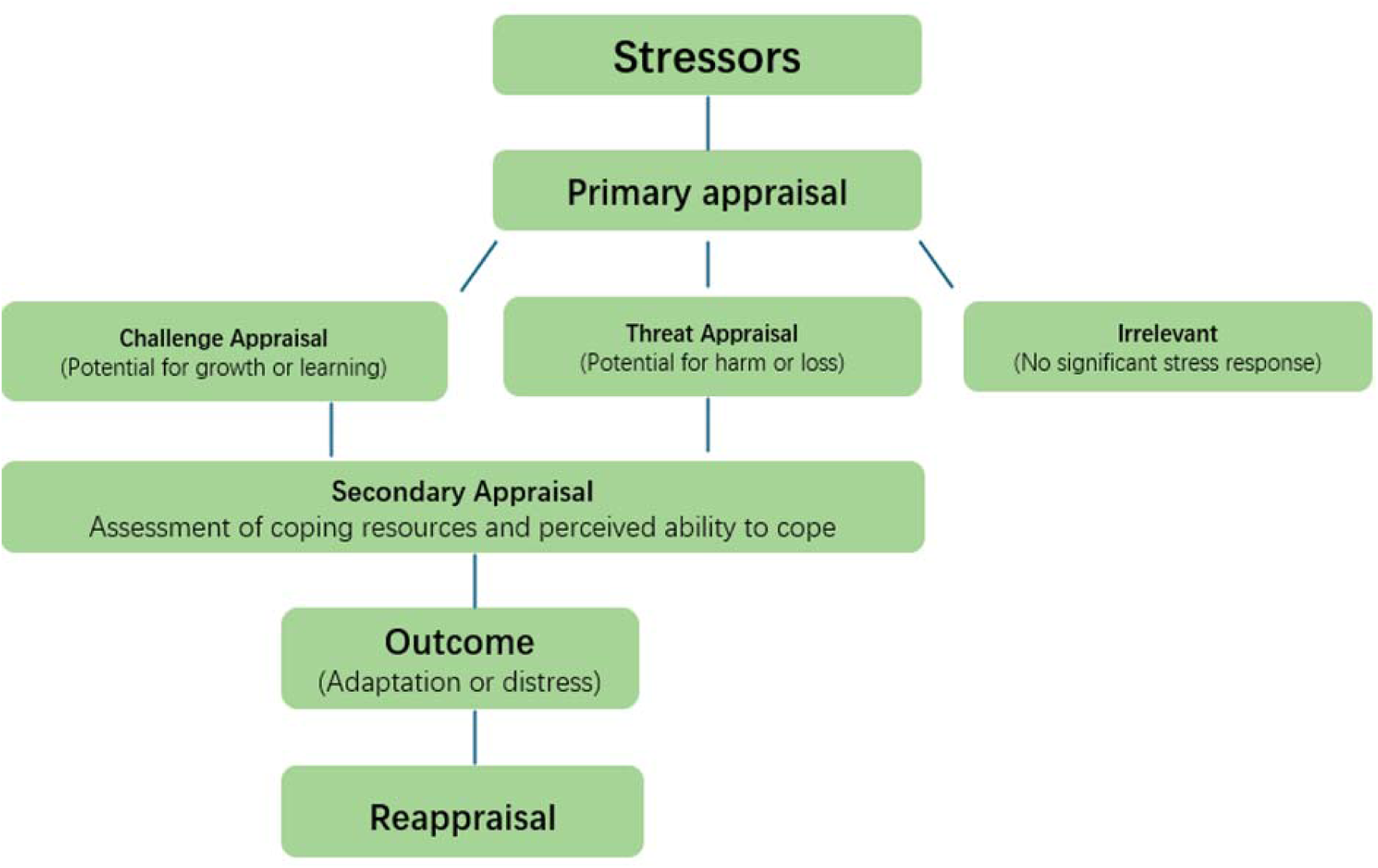
Conceptual framework illustrating the Transactional Model of Stress and Coping in nursing education.

### 2.3 Reflexivity

Reflexivity involves ongoing critical self-examination, whereby researchers consider how their assumptions and positions may influence data collection, analysis and interpretation (Gentles et al., 2014; Mitchell et al., 2018). The research team comprised of a Professor of Mental Health Nursing, a Senior Lecturer, a Lecturer, and a doctoral researcher from the School of Nursing and Midwifery, bringing complementary academic and clinical perspectives. The doctoral researcher conducted all interviews and led data collection, initial coding and theme development. A second member of the research team independently engaged with the data and developing interpretations, while the wider research team contributed to regular analytic discussions. These discussions were used to reflect on assumptions, challenge interpretations and refine the developing themes. Open-ended interview questions were used to facilitate participant-led accounts and avoid leading questions (Cairns-Lee et al., 2022).

### 2.4 Sampling

Purposive sampling was used to recruit nursing students and educators with varied experiences of stress across academic and clinical settings. Study information was disseminated via the online learning platform Moodle, email invitations, and posters with a QR code displayed in common areas within the School of Nursing and Midwifery. Students were eligible if they were aged 18 years or older, English-speaking, and enrolled in Year 2 or 3 of the Bachelor of Nursing programme; first-year students were excluded due to limited clinical exposure. Educators were eligible if they were academic or clinical staff involved in teaching or supervising undergraduate nursing students. The sample comprised six undergraduate nursing students and six nursing educators (Tables 1 and 2). Participants varied in age, gender, ethnicity and role. Students were aged 18–40 years and had all completed clinical placements. Educators included senior lecturers, a lecturer, and clinical academics.

**Table 1.** Characteristics of Study Participants – Nursing Students.

| Demographic characteristic | N = 6 |
| --- | --- |
| Gender n (%) |  |
| Female | 6 (100.0) |
| Male | 0 (0) |
| Age (years) n (%) |  |
| 18–20 | 3 (50.0) |
| 21–25 | 1 (16.7) |
| 26–30 | 0 (0) |
| 31–35 | 1 (16.7) |
| 36–40 | 1 (16.7) |
| Ethnicity<sup>a</sup> n (%) |  |
| NZ European | 4 (66.7) |
| Māori | 1 (16.7) |
| Asian | 1 (16.7) |
| Year of study n (%) |  |
| Second year | 1 (16.7) |
| Third year | 5 (83.3) |
| Prior caregiving experience n (%) |  |
| Yes | 6 (100.0) |
| No | 0 (0) |

<sup>a</sup>Ethnicity categories are reported using broad classification groupings to minimise the risk of participant re-identification.

**Table 2.**
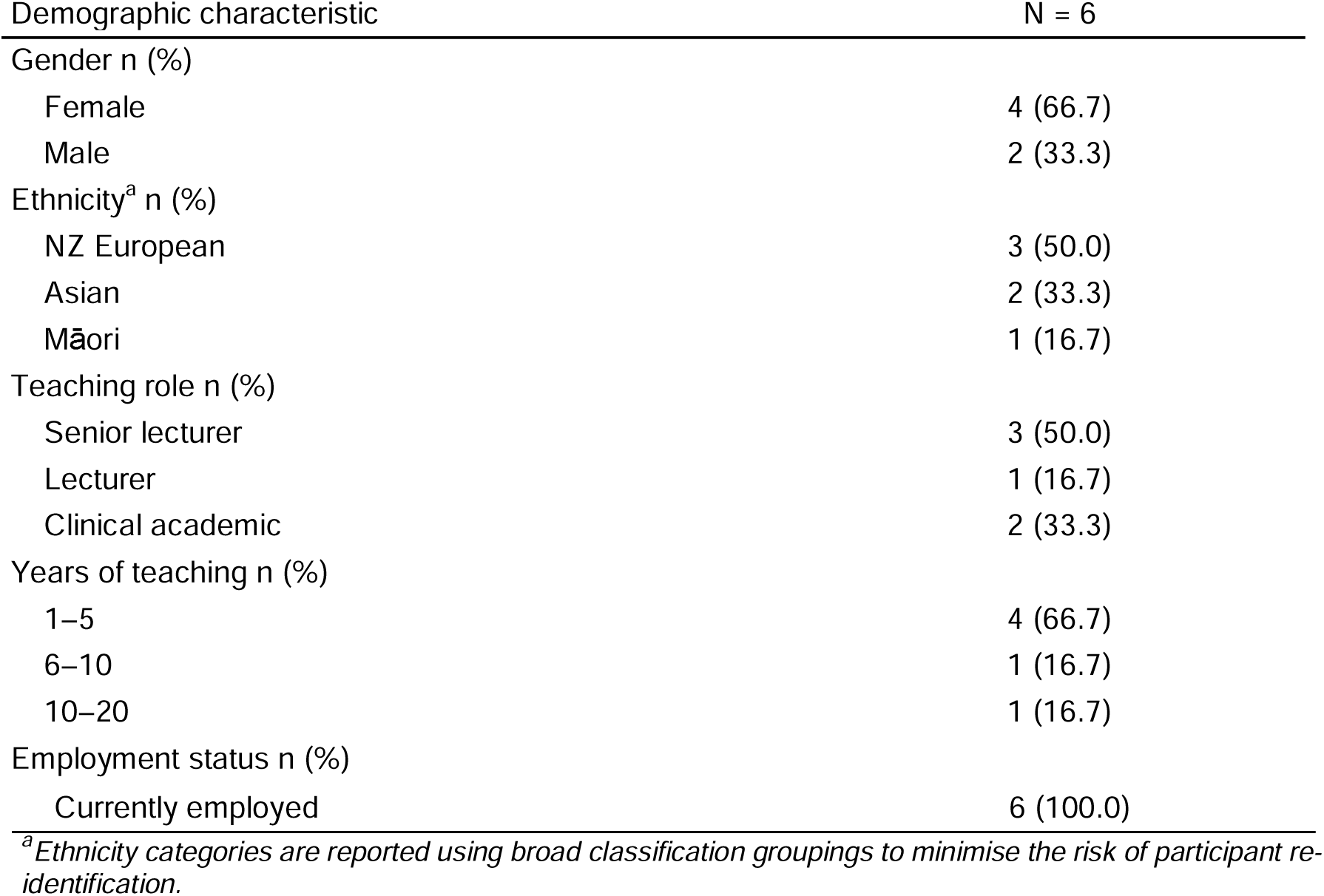
Characteristics of Study Participants – Educators.

### 2.5 Study setting

The Bachelor of Nursing programme at (xxx) university in New Zealand is a three-year, full-time pre-registration programme that meets the Nursing Council of New Zealand requirement of at least 1,000 hours of supervised clinical practice. In practice, students complete substantially more clinical hours, exceeding 1,700 hours of placement (Nursing Council of New Zealand, 2025).

### 2.6 Data collection

Data were collected between March and July 2025. Semi-structured interviews were conducted in private campus rooms or online via Microsoft Teams, with only the participant and researcher present. Interview guides with open-ended questions were developed by the research team and pilot-tested with one nurse educator. Interviews were audio-recorded with consent, lasted 30–60 minutes, and were transcribed verbatim. Transcripts were returned to participants for comment and correction.

Sample size of 12 was guided by the concept of information power (Malterud et al., 2016), whereby adequacy is considered in relation to the study aim, sample specificity, quality of interview dialogue, theoretical grounding and analytic approach rather than by a predetermined numerical threshold. The participants had direct experience of the academic and clinical contexts under investigation, and the interviews generated detailed accounts relevant to the study aim. The Transactional Model of Stress and Coping provided a sensitising conceptual framework for interpreting these accounts.

Participants were offered the option to choose their own identifiers; all consented to researcher-assigned identifiers for reporting (Wang et al., 2024). No participants withdrew from the study. Each participant was interviewed once, and repeat interviews were not conducted.

### 2.7 Data analysis

Data were analysed inductively using reflexive thematic analysis (Braun & Clarke, 2006). The (XX) led the coding process and development of candidate themes. (XX) engaged independently with the data and developing interpretations to provide an additional analytic perspective. Codes and themes were discussed within the wider research team and refined iteratively (Creswell & Poth, 2016). NVivo 15 software was used to support systematic data management.

Rigour was supported using Lincoln and Guba’s framework (Lincoln & Guba, 1985). Credibility was enhanced through in-depth engagement with the data, and regular analytic discussions within the research team to examine and refine interpretations. Dependability and confirmability were addressed through reflexive consideration of the analytic process, further supported by supervisory review.

### 2.8 Ethical considerations

Ethical approval was granted by the Human Research Ethics Committee Number xxxx). Participation was voluntary and confidential; recruitment notices were distributed through institutional channels, and the doctoral researcher contacted only those who expressed interest, thereby minimising potential coercion. Informed consent was obtained prior to participation. Members of the research team who held teaching roles had no direct relationship with participants and no access to identifiable data; all transcripts were de-identified. Cultural consultation was undertaken in accordance with institutional research ethics processes to ensure cultural appropriateness and respect for Māori perspectives.

## Findings

Analysis generated three themes and eleven subthemes describing nursing students’ and educators’ perceptions of stress, its impacts, stress appraisal and coping, and supports and barriers across academic and clinical contexts (Figure 2).

**Figure 2:**
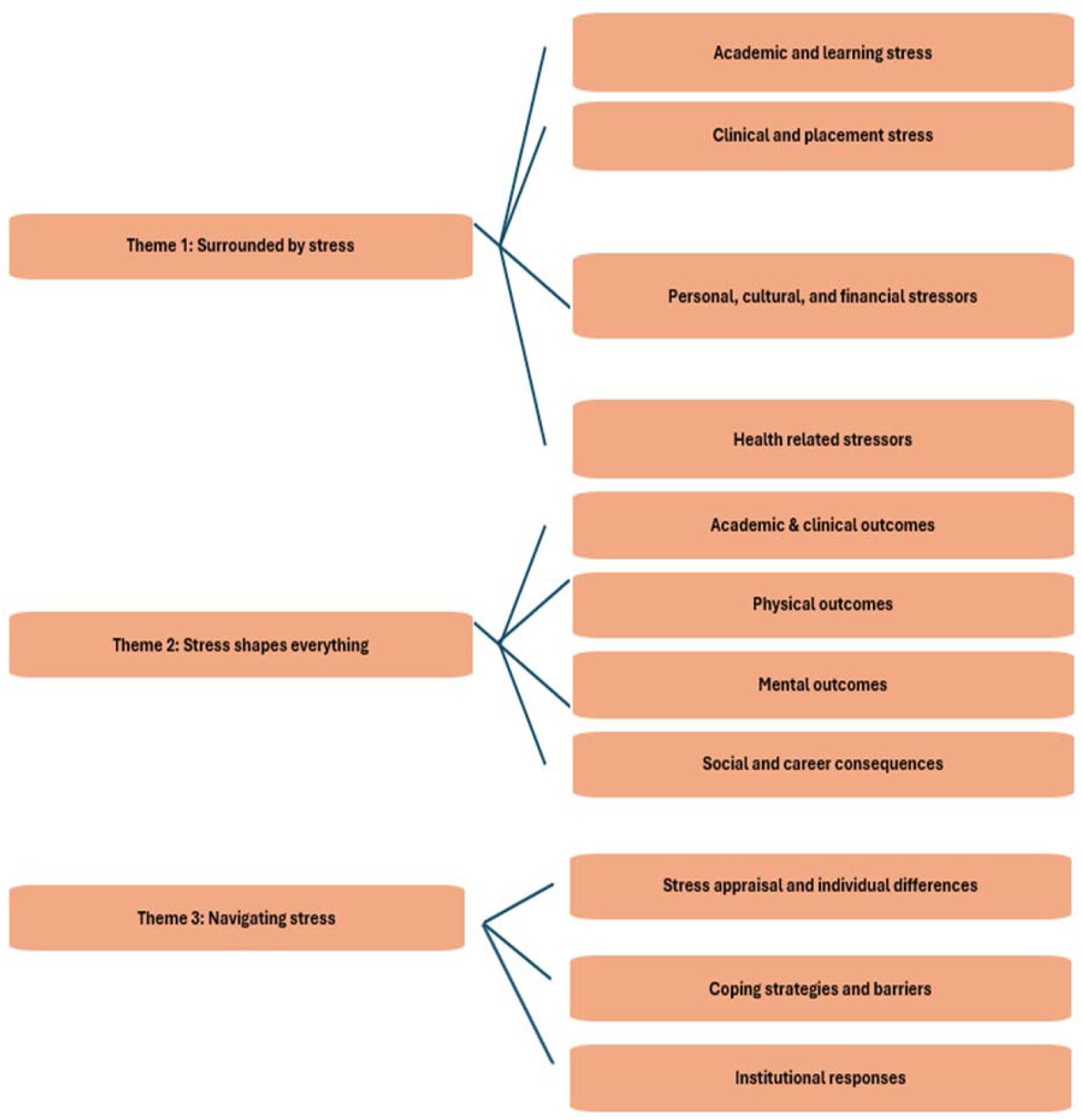
Themes and subthemes

Participants are identified as S1–S6 (students) and E7–E12 (educators). Coded segments reflect contributions from all participants, and this system was used consistently to ensure clarity and anonymity.

## Theme 1: Surrounded by stress

This theme explores situations perceived to contribute to undergraduate nursing students’ stress. Four main sources were identified: academic demands, clinical placement experiences, personal and social factors, and health-related stressors.

Subtheme 1: Academic and learning stress

Academic expectations were identified as a major source of stress. Students described placing high pressure on themselves, often linked to perfectionism, fear of failure, and self-doubt regarding academic adequacy.

> “I’m quite a perfectionist, and I put a lot of pressure on myself with my academics, and so I’m always going to be stressed…” (S3)
>
> “the stress … good and bad, but for myself bad, because I’m… hard on myself. Like, did I do enough? Did I do this? Did I do that?” (S6)

Educators noted that family expectations, particularly for international students, could intensify academic pressure.

> “A lot of international students have quite an expectation from the family point of view… you have to be…an A student “(E7)

Students also reported stress related to perceived knowledge gaps, particularly after time away from academic study during placements and around examinations.

Educators did not raise this concern, suggesting a gap between students’ self-perceived academic adequacy and educators’ recognition of this stressor.

> “As soon as we come back from placement… we got a couple of tests… you got to study for this test… once you go back to (class).” (S1)
>
> “I did stress… around the academic side, just because it had been so long [from placement] since I had done anything academic…” (S2)

Academic workload was perceived as a significant source of stress, particularly the need to balance academic requirements alongside clinical placements. Students described assignment clustering and tight deadlines as especially demanding.

> “We’ve got one assignment due this Friday, and then two assignments due… and then another due next Tuesday…” (S1)

Tests scheduled soon after placements were also reported as stressful. Educators acknowledged these pressures but framed them as an inherent aspect of professional preparation, emphasising the need for students to develop time-management skills.

> “…if you work in clinical area, you are not going to have luxury of time writing your assignment. That means you have to be super organised.” (E12)

The transition to the tertiary learning environment was described as challenging. Educators identified expectations of self-directed learning as a key stressor.

> “…all of a sudden you’re in this university situation, where you actually [have] to plan what you want to learn.” (E10)

However, students did not explicitly label this transition as stressful, instead referring to difficulties such as pressures of group work.

> “A group stresses me out a lot… you can feel the vibe of the group.” (S1)

Both students and educators also noted stress from unfamiliar situations, including new learning systems and placements.

> “…it was going into uni…then out in placement… It was something I hadn’t really experienced before.” (S4) “In their first year they’re[students] just like scared rabbits… not quite sure what’s happening.” (E9)

Subtheme 2: Clinical & Placement Stress

Participants disclosed stress in clinical contexts. Both students and educators described that stress usually came from a demanding clinical workload and from having to juggle placement demands alongside ongoing academic tasks.

> “I used to come over [home] from placement. I’d be really tired; I couldn’t do anything [academic work] after placement. ” (S4)
>
> “For our program… we have very high clinical hours compared to some other programs, so students spend relatively longer times in the clinical area…” (E10)

Beyond workload, students and educators also linked stress to the emotional demands of clinical work, including witnessing patients’ suffering and fear of making mistakes or causing harm.

> “… very stressful, stressful environment to work in, because you’re working with…people who are sick, obviously, that are challenged themselves[students].” (E11)
>
> “…being worried that you’re going to do something that’s going to harm someone[patients] instead of helping them.” (S3)

Attitudes of preceptors was also reported as a stressor. From students’ perspectives, unwelcoming preceptors heightened stress, whereas positive preceptors fostered learning.

> “…if the preceptors… they’re welcoming… I would have more of a mindset to learn. But if, if [the] preceptor is… not very welcoming… kind of ignore you, that would stress me out, and that would not put me in a learning mode.” (S1).

One Māori (Indigenous peoples of Aotearoa New Zealand) student reported additional stress when repeatedly being assigned Māori patients during placement. This was perceived as cultural stereotyping and racism and reflected broader structural issues within the clinical learning environment.

> “I was given the people of my ethnicity, which was cool… but I’m still the student… I am still learning… you’re pushing me towards my own ethnicity” (S6).

Subtheme 3: Personal and Social Stress

All participants described stress arising from personal and social factors beyond academic and clinical demands. Financial pressure was a prominent stressor, with students struggling to balance study costs, living expenses and part-time work.

> “I just put all my thing on my student loan, and then work during the…holidays” (S4)
>
> “…a lot of them[students] are juggling part time jobs… there’s kind of that stress of money.” (E7)

Educators noted that students’ also experienced stress from balancing family responsibilities, with little time for rest.

> “Those that are parents moving into study and then balancing their parental responsibilities… that’s one of the stresses… doing the student life but also making sure family life is prioritised.” (E7)

For international students, stress was intensified by family separation, cultural adjustment and language barriers, compounded by limited support networks.

> “Some of the overseas students [that] English is their second language. They’ve got no support system around them… different country, different language …students that become isolated in their family life… their disconnection from everything.” (E9)

Final-year students additionally reported anxiety related to future employability, describing their last year as a continuous assessment of readiness for future work.

> “…in third year…this whole year is like a job interview and so being worried that you’re going to do something that impedes your chances of getting a job” (S3)

Subtheme 4: Health-related stress

Educators and students were aligned in describing how chronic illness and fatigue disrupted students’ focus, attendance, and ability to complete coursework.

> “[student] has chronic health issues… might not be present for this lecture…have to travel… to other area for specialist review.” (E10)
>
> “I have chronic illness and so I have less time and less energy… I have the same amount to do, but not as much time or energy.” (S3)

## Theme 2: Stress shapes everything

This theme explored the range of academic, clinical, physical, and psychological consequences including relationships and career considerations associated with stress.

Subtheme 1: Academic and clinical outcomes

Participants reported that stress significantly affected students’ engagement, motivation, concentration and performance.

> “If I get stressed, I can’t really focus on my uni work.” (S4)
>
> “I don’t want to study because then I’ll get stressed, and then I just put off studying until the very, very last minute.” (S5)

Educators identified disengagement as the most visible response. In more severe cases, sustained stress contributed to students leaving the programme entirely.

> “they [students] drop out. They withdraw… they’ll disengage.” (E11)

Subtheme 2: Physical consequences

Students and educators were aligned in observing that stress was not only felt emotionally but was also visible in students’ physical health, for example acne.

> “I found last year was quite stressful… I ended up getting really bad acne” (S2)

Sleep disruption was one of the most reported symptoms. Students described lying awake ruminating over coursework, struggling to recover after late shifts, and finding it difficult to maintain a healthy routine. Educators echoed these observations, noting that poor sleep reduced concentration.

> “Sleep… it ticks over my head when I try to have good sleep hygiene… all my final papers are coming in my head.” (S6)
>
> “if you don’t have good sleep, It’s really hard to sit in class and listen …not being able to follow the content…” (E8)

In severe cases, stress escalated into maladaptive coping, with some students turning to substances to escape pressure or maintain functioning.

> “some students will migrate to using substances like alcohol or illicit drugs or things to try to keep them going…” (E8)

Subtheme 3: Psychological consequences

Students and educators described stress as contributing to mental health concerns, particularly anxiety, burnout, and overthinking. Students commonly experienced anxiety as an immediate and ongoing response to stress, alongside feeling unsettled or unable to manage worries.

> “… I wasn’t like, you know, like feeling good in myself…that was…from stress…I’d…get anxious” (S2)

Stress also affected everyday behaviours, with students describing reduced engagement in self-care when overwhelmed.

> “If I’m stressed… I don’t want to… put cream on my face… I completely ignore that part … all the dressing things and everything.” (S1)

In contrast, educators noted that recognising stress symptoms could represent a positive step toward self-regulation and help-seeking.

> “I had one student last week… saying…I’m really fatigued. I need to go home and rest. And that’s a really good sign, because the student actually recognised the signs for stress.” (E10)

Subtheme 4: Social and career consequences

Stress extended beyond academic, clinical, physical, and psychological domains, influencing students’ relationships and future career considerations. Students described withdrawing socially when stressed, prioritising workload over connection with peers and family.

> “When I’m stressed… I usually don’t talk to them… I just want to be by myself and then get the stuff done.” (S1)

Educators observed that ongoing stress could lead some students to lower their career ambitions to avoid future pressure.

> “they [students] may even take up a role where that level of education is not required.” (E11)

## Theme 3: Navigating stress

Students and educators described stress as a dynamic experience shaped by individual appraisal, coping responses, and the broader educational context.

Participants highlighted differences in how stress was understood and managed, as well as structural factors that supported or constrained effective coping.

Subtheme 1: Stress appraisal and individual differences

Stress was viewed as having a dual effect, described as both overwhelming and motivating. One educator noted that students primarily associated stress with negative outcomes and rarely viewed it as beneficial.

> “In the students’ brain, they don’t really see stress as a positive thing… I haven’t had any students say, I love this stress…” (E10)

However, some students and educators described stress as motivating, helping students maintain focus or support learning when psychological safety was present.

> “I try to use stress to motivate me a bit… and try and take that and do something about it.” (S3)
>
> “Sometimes stress makes me remember things better… I guess stress is sort of a positive factor for me when it comes to academic things.” (S1)
>
> “Some stress is good… if people are feeling a little bit challenged but still feel psychologically safe, they are going to be more open to learning.” (E11)

Educators emphasised that stress responses were highly individual and shaped by previous experiences and available support.

> “It is very individualistic… if the student has not experienced the level of stress they experience, they’re going to be acting differently. Then it depends on your support system… So that all makes difference.” (E12).

Subtheme 2: Coping strategies and barriers

Participants described a range of coping strategies that supported wellbeing. Peer support was frequently identified as a key resource that reduced isolation and provided emotional reassurance.

> “It was really helpful for them [students] to get together to share their experiences… the peer support” (E11).
>
> “Peers… have supported me… I’m very grateful for that too, because you could be feeling isolated…being 20, 30 minutes’ drive away from [campus].” (S6).

However, peer dynamics could also have negative effects, as comparisons with peers sometimes heightened stress.

> “I find myself stressed… comparing our content with…friends who study at other universities … Sometimes… you feel…they’re more ahead than the content that they’ve been that’s been taught to us” (S1).

Students also described using physical activity, mindfulness, and cultural or spiritual practices to regulate stress. Exercise, walking, and sport helped restore focus, while prayer, breathing, and self-care promoted relaxation.

> “I do a lot of walks in the evening… that is a big de-stressor for me.” (S1)
>
> “I pray every day…of our own religion. And that settles me for the day too” (S6)

Support from lecturers was highly valued by both groups, as approachable staff offered care that eased academic and emotional pressure.

> “I always felt like I could go and talk to a lecturer… I knew that there would be support for me… All the lecturers are really good… warm and welcoming.” (S2)
>
> “My view is as a lecturer trying to build a good, positive relationship with students leaving the door open… if [students] find your lecture is accessible… they find it much easier to deal with it…” (E12)

However, some students perceived lecturer–student relationships as too formal, which created hesitation in seeking help. Educators similarly noted that students often disclosed difficulties only after direct prompting.

> “[Even] If I want to approach them [educators], I won’t [try to talk to educators], just because they’re my lecturer…” (S1).
>
> “You don’t know why this person [student] has not been engaged in any online activities, and when you question them, they will just say… actually I’ve been feeling really unloved for two weeks, but there’s no disclosure” (E10).
>
> “They perceive their lecturers very busy. They are very hesitant to contact them [educators].” (E12)

Additionally, family and partners also offered motivation and understanding for students. Taking recovery time, whether short breaks or full days off, was viewed as essential for maintaining focus and preventing burnout.

> “…my partner… He’s receiving it, so I guess he gets the best, worst part of [stress]’” (S1).
>
> “Give themselves [students] a break… they’ll be able to respond after they’ve centred themselves, rather than react to something…” (E7)

Subtheme 3: Institutional responses

University-based resources, particularly counselling and health services, were identified as important supports for managing stress.

> “If you [students] psychologically or physically have something happening, go to the health centre… it’s all covered [by student fees and health subsidies].” (E12)

However, experiences of accessing support were mixed. One student reported receiving general advice (e.g., sleep hygiene) from the health centre but felt their concerns were not considered severe enough to warrant in-person mental health support, which they experienced as frustrating.

> “I contacted them [mental health services] on the phone… I don’t think they had many… in person appointments available… So, they did a phone call… I felt like my problem… wasn’t bad enough… I just felt…let down… it wasn’t … a top priority.” (S2).

Similarly, one educator observed that some students lacked awareness of the services available to them, suggesting a need for greater visibility and accessibility.

> “Not visible enough [mental health service] … And even as a lecturer, we’re not quite clear… the process isn’t very transparent” (E10).

Participants also valued person-centred programme cultures, wellness initiatives, and environmental features such as quiet or green spaces that supported relaxation.

> “Our [nursing] program is quite person-centred… we’ve put in a Wellness Week… not just focused on the academic but on the person.” (E7)
>
> “Having quiet spaces where people can zone out… green space… some environmental factors contributing to de-stress.” (E8)

Several areas for improvement were highlighted, including earlier help-seeking, nursing-specific services, and culturally responsive support, particularly for international and Pacific students, while maintaining inclusive approaches for all learners.

> “Maybe have something like dedicated to nursing students, because we don’t see much resources… it’s just for the general university student.” (S1).
>
> “…providing specific support for our Asian students or international students in general, especially people who speak English as a second language.” (E10).

In contrast with the need for culturally specific support one participant reported the need for services that were inclusive of all students.

> “I’ve seen that separation, like a Māori stream and a mainstream. I actually do like the involvement of all… more inclusive for all…” (S6).

Broader wellbeing initiatives extending beyond academic concerns were suggested, including practical supports such as improved access to free parking. Educators also noted that systemic constraints, particularly workload pressures, limited their capacity to provide adequate student support.

> “I’ll do [suggest university] that [to provide more] parking and no cost” (S6)
>
> “Leadership [needs] to make sure… they are attending to the wellness of each student, not just leaving it to us[educators].” (E7).

## Discussion

This study provides novel insights into stress by simultaneously exploring perceptions of stress among undergraduate nursing students and educators within the same programme. Bringing these perspectives together highlights the multiple, intersecting sources of student stress and important differences in how stress is perceived and responded to within nursing education.

### Surrounded by stress

Academic demands emerged as a prominent source of stress, including workload intensity, assessment pressure, and fear of failure. While academic workload and performance expectations are widely recognised stressors in nursing education (Dasgupta et al., 2020; Labrague et al., 2017), the findings of this study suggest that these pressures are not only externally imposed but also internally reinforced through students’ high self-expectations and academic insecurity. This was particularly evident during clinical placement periods, when academic responsibilities continued alongside demanding practice environments.

A notable difference between student and educator perspectives was evident in how academic stress was interpreted. Students often described feeling overwhelmed by expectations and uncertainty about their academic performance, whereas educators tended to frame stress as an inherent component of professional development. This divergence highlights a potential perception gap within nursing education, where educators may view stress as a developmental challenge while students experience it as a source of persistent pressure (Dogham et al., 2024; Zeb et al., 2022). Stress related to group work and unfamiliar learning environments reflected the social and cultural demands of nursing education. Previous researchers have shown that collaborative learning can increase stress among students, while international students may experience additional stress when adjusting to new cultural and language contexts (Graham et al., 2024; Mao et al., 2023).

Stress associated with clinical placement has been consistently reported in nursing education research (Admi et al., 2018; Ahmed & Mohammed, 2019). In this study, clinical placements were reported by students and educators as a major stressor, particularly when combined with ongoing academic demands. Students described fatigue, emotional strain, and anxiety related to patient safety, consistent with evidence linking higher clinical hours to increased stress and burnout risk (Mohamed et al., 2024). Interpersonal dynamics within clinical environments also shaped stress experiences. Preceptor attitudes were particularly influential, with supportive supervision fostering confidence and learning, while unwelcoming behaviours increased anxiety and reduced perceived learning opportunities (Vellem & Jooste, 2025). Fear of harming patients or witnessing suffering was salient when students perceived limited competence or control, reflecting findings that identify patient safety concerns as a key source of clinical stress (Ching et al., 2020). In addition, cultural and relational dynamics, including experiences of stereotyping and racism, further intensified stress, highlighting the importance of culturally safe and inclusive supervision for ethnically minoritised nursing students in New Zealand (Hunter & Cook, 2024; Walker et al., 2024).

Participants in this study reported that financial strain, part-time work, and family responsibilities contributed to fatigue and limited opportunities for recovery, reflecting findings in previous research that identify these factors as key stressors among nursing students (Andrew et al., 2022; Labrague et al., 2017).

### Stress shapes everything

Students commonly reported reduced motivation, concentration, and engagement, with some describing avoidance behaviours and burnout during high-pressure periods. These experiences suggest that stress may disrupt students’ engagement with learning, contributing to academic disengagement reported in previous research (Sheikoleslami et al., 2025). Physiological symptoms such as fatigue and sleep disturbance reflected the effects of stress observed in other nursing student populations (Ibrahim et al., 2024). Neglect of self-care and increased substance use were also consistent with maladaptive coping patterns reported in previous research (Dias et al., 2024). Anxiety and burnout were particularly evident during high-pressure periods, aligning with previous studies demonstrating that stress is positively correlated with anxiety and burnout (Hwang & Kim, 2022; Onieva-Zafra et al., 2020). However, reflecting differences between student and educator perspectives, one educator suggested that recognising stress may represent an early step toward help-seeking and adaptation. This view aligns with research indicating that awareness of stress can facilitate adaptive coping and engagement with support strategies (Zeb et al., 2022).

### Navigating stress

Divergent perceptions were reported from participants, with some students describing stress as motivating and performance-enhancing, while most experienced it as overwhelming and demotivating. This suggests that stress is not a uniform experience but is shaped by individual appraisal and coping capacity, which is consistent with Lazarus and Folkman’s transactional model of stress and coping (Lazarus & Folkman, 1984). Previous research similarly indicates that moderate stress may function as eustress, supporting engagement and resilience, whereas excessive stress undermines wellbeing (Fonseca et al., 2019). However, research examining positive stress among nursing students remains limited, and most participants in this study associated stress with negative emotional outcomes, including anxiety and exhaustion (Watson et al., 2025).

Nursing students used multiple strategies to manage stress, with peer support emerging as a key coping resource that reduced isolation and provided reassurance (Harvey et al., 2024). However, peer interactions also involved comparison, which could intensify stress (Kwak et al., 2022). Institutional supports, including counselling, wellness initiatives, were valued but inconsistently accessed, as students were often unsure of service availability or perceived their concerns as insufficiently serious.

Educators also play an important role in fostering a culture in which stress can be shared and in supporting students’ stress management (Zeb et al., 2022) . Educators’ approachability further influenced help-seeking, with empathetic and flexible staff fostering psychological safety, while more formal relationships discouraged disclosure (Froneman et al., 2016). Self-care practices, including exercise, mindfulness, and prayer, supported focus and emotional balance, aligning with evidence highlighting their protective role in nursing student wellbeing (Rafati et al., 2017; Veigh et al., 2021).

Both students and educators identified the need for nursing-specific and culturally responsive support, reinforcing literature emphasising culturally safe environments for student wellbeing and retention (L. Labrague et al., 2018; Nursing Council of New Zealand, 2011; Solomon et al., 2025). References to practical supports, such as accessible parking, further illustrate that student wellbeing extends beyond academic concerns and requires a shared institutional response (New Zealand Nurses Organisation, 2023).

### Towards a transactional model of stress and coping in nursing

The findings suggest that stress among nursing students is a dynamic and contextual process rather than a uniform experience. Previous studies have applied Lazarus and Folkman’s transactional theory of stress and coping to understand undergraduate nursing stress, particularly within clinical settings. A qualitative study has explored coping strategies used during clinical placements, highlighting reflection, social support, and self-awareness(Kaur et al., 2020), while quantitative research has examined relationships between personal resources such as self-efficacy and placement stress(Kim et al., 2019). Conceptual applications in mental health nursing have further emphasised appraisal processes and emotion regulation in challenging clinical contexts (Martin & Daniels, 2014).

Building on this work, our findings indicate that stress extends beyond isolated practicum experiences and occurs across academic, clinical, and personal domains. Drawing on the widespread nature of stress across academic and clinical environments (Theme 1) and its multiple impacts (Theme 2), students demonstrated varied appraisals of demands and diverse coping responses (Theme 3). Different students interpreted similar situations differently and selected strategies they perceived as effective. This supports a transactional understanding of stress and coping, whereby outcomes are shaped by ongoing appraisal, individual interpretation, and available resources (Lazarus & Folkman, 1984).

These findings suggest that early experiences of stress, coping, and emotional demands may shape mental wellbeing, resilience, and vulnerability to distress in the future nursing workforce. The findings highlight the importance of early identification of stress, culturally responsive learning environments, and targeted supports that promote adaptive coping across academic and clinical contexts.

### Strengths and limitations

This study provides an in-depth qualitative exploration of stress integrating the perspectives of undergraduate nursing students and educators within the same programme, something that has not been previously reported. The study identified both similarities and differences in the perceptions of each group. As a single-institution study, transferability may be limited, and reliance on voluntary self-reported data may introduce bias. Nevertheless, the findings align with existing research and offer context-specific insights to inform mental health support initiatives in New Zealand nursing education.

## Conclusion

This study explored how nursing students and educators in New Zealand perceive stress and coping across academic and clinical settings. Stress was understood as both positive and negative, with coping shaped by personal, social, and institutional factors. The findings support a transactional understanding of stress, highlighting its dynamic and context-dependent nature. Early patterns of stress appraisal and coping may shape mental wellbeing, emotional regulation, and vulnerability to distress in the future workforce. Effective support therefore requires flexible, culturally safe, and institution-wide approaches that promote mental wellbeing and strengthen adaptive coping.

## Data Availability

The interview data generated and analysed during the current study are not publicly available due to ethical and confidentiality restrictions.

## Notes

### Competing Interest Statement

The authors have declared no competing interest.

### Author Declarations

The Human Research Ethics Committee (Health) of the University of Waikato gave ethical approval for this work (HREC(Health)2025#06).

